# An economic evaluation of two self-sampling strategies for HPV primary cervical cancer screening compared with clinician-collected sampling

**DOI:** 10.1101/2022.11.03.22281845

**Authors:** Susie Huntington, Krishnan Puri Sudhir, Verena Schneider, Alex Sargent, Katy Turner, Emma J Crosbie, Elisabeth J Adams

**Affiliations:** Aquarius Population Health, Unit 29 Tileyard Studios, London N7 9AH, UK; Cytology Department, Clinical Sciences Centre, Manchester University NHS Foundation Trust, Oxford Road, Manchester M13 9WL, UK; Division of Cancer Sciences, Faculty of Biology, Medicine and Health, University of Manchester, Oxford Road, Manchester M13 9WL, UK; Department of Obstetrics and Gynaecology, St Mary’s Hospital, Manchester University NHS Foundation Trust, Manchester Academic Health Science Centre, Oxford Road, Manchester M13 9WL, UK

**Keywords:** HPV infection, cancer screening, economic evaluation, self-sampling

## Abstract

**Objective:** To compare the cost and effects of three sampling strategies for human papillomavirus (HPV) primary screening.

**Design:** Cost-consequence analysis using a decision tree in Excel.

**Setting:** England.

**Participants:** A cohort of 10,000 women age 25 to 65 eligible for the NHS Cervical Screening Programme (NHSCSP) (Box 1).

**Methods:** The model was informed by the NHSCSP HPV primary screening pathway and adapted for self-sampling. It used a 3-year recall cycle with routine screening in year 1 and recall screening in years 2/3. Parameters were obtained from published studies, manufacturers, NHSCSP reports, and input from experts.

**Interventions:** Three sampling strategies were: 1) routine clinician-collected cervical sample, 2) self-collected first-void (FV) urine; 3) self-collected vaginal swab. The hypothetical self-sampling strategies involved women being mailed a sampling kit at home.

**Main outcome measures:** Primary outcomes: overall costs (for all screening steps to colposcopy), number of complete screens, and cost per complete screen. Secondary outcomes: number of women screened, number of women lost to follow-up, cost per colposcopy, and total screening costs for a plausible range of uptake scenarios.

**Results:** In the base case, the average cost per complete screen was £56.81 for clinician-collected cervical sampling, £38.57 for FV urine self-sampling, and £40.37 for vaginal self-sampling. In deterministic sensitivity analysis (DSA), the variables most affecting the average cost per screen were the cost of sample collection for clinician-collected sampling and the cost of laboratory HPV testing for the self-sampling strategies. Scaled to consider routine screening in England, if uptake in non-attenders increased by 15% and 50% of current screeners converted to self-sampling, the NHSCSP would save £19.2 million (FV urine) or £16.5 million (vaginal) per year.

**Conclusion:** Self-sampling could provide a less costly alternative to clinician-collected sampling for routine HPV primary screening and offers opportunities to expand the reach of cervical screening to under-screened women.

**Strengths and limitations of the study:**

- This is the first study to assess the cost of screening for cervical cancer using self-collected first-void urine or vaginal swab compared to the current strategy of clinician-collected cervical sampling within the context of England’s NHS Cervical Screening Programme (NHSCSP).
- The cost per screen was used to calculate the total cost of the NHSSCP in England, allowing a comparison of different uptake scenarios if self-sampling was offered to non-attenders only or to all eligible women.
- Limited published data were available to inform the cost of self-sampling devices and HPV laboratory testing of self-collected samples.
- One pathway for self-sampling was examined. However, there are alternative pathways which could be explored, some of which are dependent on new technologies, such as DNA methylation testing, being validated and costed.

## Introduction

Cervical cancer is a leading cause of mortality among women worldwide but can be prevented through screening for high-risk human papillomavirus (HR-HPV) [1–3]. HPV positive samples are triaged by cytology to identify women at risk of high-grade cervical intraepithelial neoplasia (CIN2+) and treat them to prevent progression to cancer. The impact of cervical screening programmes is heavily dependent on achieving high participation rates [4]. Screening uptake in England fell from 76% in 2010/11 to 72% in 2019/20 decreasing further during the COVID-19 pandemic, to 70% in 2020/21 when there were disruptions to the health service and in-person appointments were minimised [4–6].

In England and many other countries, screening requires attendance at a healthcare facility where a healthcare professional collects a cervical sample [7]. Reported reasons for non-attendance include difficulty making an appointment, embarrassment, fear, and inconvenience [8,9]. There has been a growing interest in the use of vaginal self-sampling for HR-HPV testing due to its relative convenience and acceptability among women [10–13]. Home self-sampling is successfully used in other screening programmes e.g., bowel cancer [14] and chlamydia [15] and for some other sexually transmitted infections [16]. Countries including Australia, Denmark, Malaysia, and The Netherlands, have or are moving to self-sampling as a screening option [17–19]. In 2021, NHS England launched YouScreen, a pilot study in targeted locations offering vaginal self-sampling kits to individuals overdue for screening [20].

Self-collected first-void (FV) urine is another option for HPV screening [10,21,22]. There is growing evidence that if HR-HPV assays are optimised for use on urine or self-collected vaginal samples their diagnostic performance is non-inferior to that achieved for clinician-collected cervical samples [23– 25]. Currently, within the NHSCSP, cytology is performed on HR-HPV positive samples to determine whether colposcopy is required. Cytology cannot be performed on urine or vaginal swab samples, so an HR-HPV positive result means a second, clinician-collected sample is required. Despite this additional step, which might be avoidable with new technologies [26], self-sampling is likely to be cost-saving and remove barriers to screening. It has the potential to increase uptake in those who do not currently screen, referred to as non-attenders, and improve user experience and choice [27,28].

This study aimed to compare the cost of cervical screening (including sample collection, HPV testing, cytology and colposcopy) using three sampling strategies: clinician-collected cervical sampling, FV urine self-sampling, and vaginal self-sampling. Scenarios were explored using different uptake rates and offering self-sampling to everyone or only to non-attenders. The results will inform decision-makers about the screening costs and effects of offering self-sampling as an alternative to routine screening in HPV primary cervical screening programmes.

## Methods

### Model type and structure

A decision tree model was constructed in Excel v2202 (Microsoft, Redmond, WA, USA) to simulate a hypothetical cohort of 10,000 people invited for screening. The model’s structure was based on the HPV primary screening algorithm for cervical cancer screening used in England (See Box 1) and many other countries (Figure 1) and based on a previous model [29,30].

**Figure 1.**
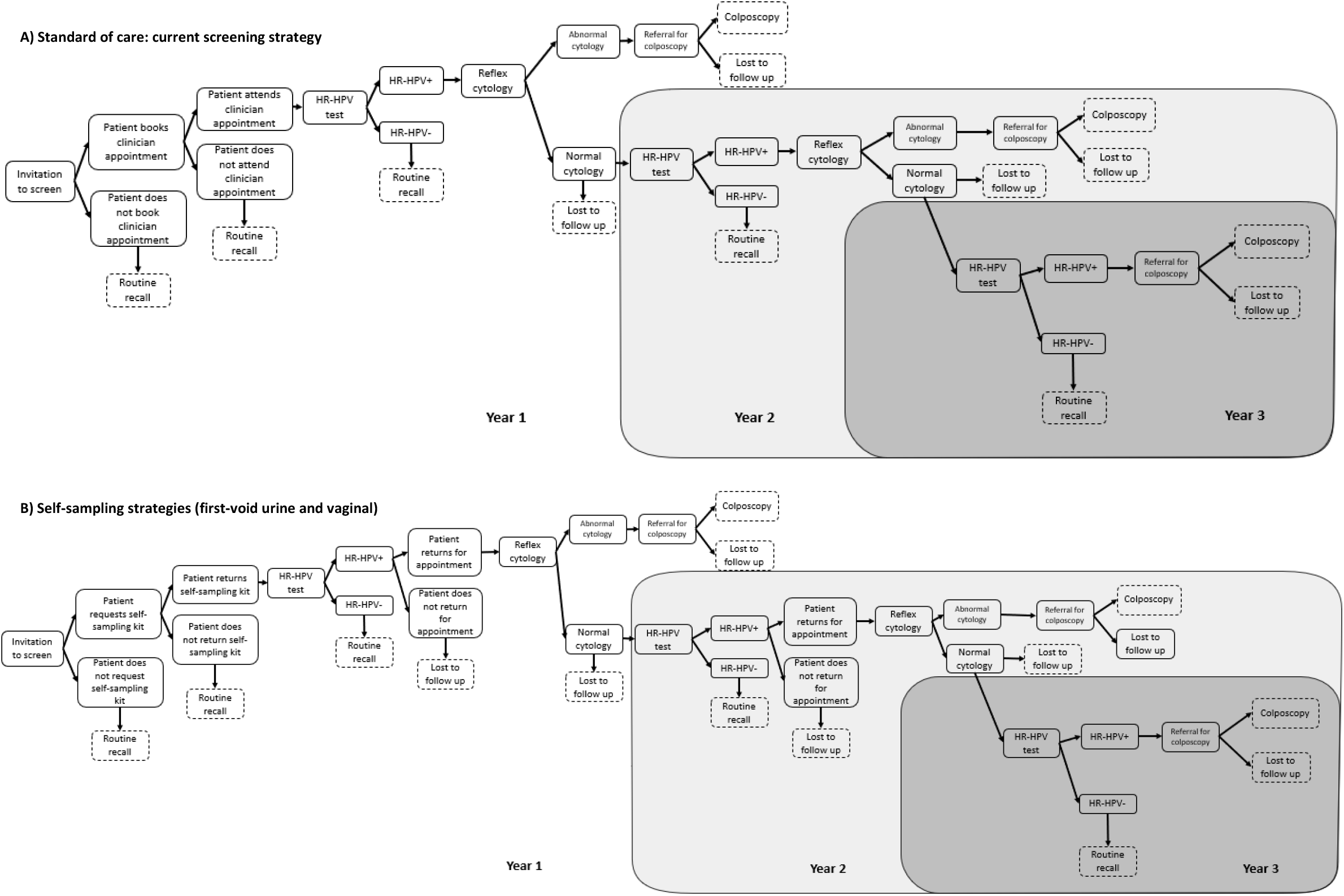
Structure of the decision tree model to simulate cervical cancer screening in England. Footnote: Women who in Year 1/2 have an HR-HPV positive result and normal cytology but do not attend for clinician-collected sampling (A) or provide a self-collected sample (B) the following year are considered lost to follow-up. The cost of non-attendance in primary care or non-return of requested sampling kits in Years 2/3 are not considered.

#### Box 1

**Screening eligibility in England’s NHSCSP**

In England, women and people with a cervix age 25 to 64 are eligible for cervical screening every 3 years (for those age 25 to 49) or every 5 years (for those age 50 to 64). Invitations to screen as part of the NHSCSP are automatically sent to anyone registered with a general practice (GP) as female. People with a cervix who are registered as male are not automatically sent an invite (due to limitations of the IT system currently used) but should receive an invite sent by their GP practice or healthcare teams managing gender reassignment and can request screening themselves.

We refer to people in our model cohort as ‘women’ reflecting the reality of the current system and acknowledging that some people with a cervix who are not registered as a woman might be missed despite being eligible for cervical screening.

### Comparators

A cost-consequence analysis was performed to compare three sampling strategies within the context of the NHSCSP: clinician-collected cervical sampling (standard of care, SoC), FV urine self-sampling and vaginal swab self-sampling. In the base case, eligibility for screening (Box 1), and screening uptake (attendance in primary care for clinician-collected sampling or return of sample by post) was the same for all strategies. It was assumed that there was equivalent HPV test sensitivity and specificity for each sample type.

The self-sampling strategies were chosen because they are acceptable to patients and are already used for home testing in other disease areas. The Colli-Pee® device (Novosanis NV, Wijnegem, Belgium) [31] collects a standardised volume of FV urine without the need for the person to interrupt the flow of urine. The use of a standard urine pot was not assessed since testing of mid/random flow urine for HPV is not as accurate as FV urine [32].

### Time horizon

The model considered a three-year time horizon. This period includes the minimum time before recall to routine screening and includes the complete cycle of events for HR-HPV positive women with normal cytology in years 1 and 2 (Figure 1). No long-term outcomes or costs associated with cancer diagnosis or treatment were considered.

### Current cervical screening pathway

At present, within the NHSCSP (Figure 1), people eligible for screening are invited by letter to attend an appointment at their GP during which a cervical sample is taken by a clinician. The sample is sent to one of eight laboratories where it is tested for HR-HPV. HR-HPV negative women are discharged to routine recall. HR-HPV positive samples are tested for abnormal cell changes using liquid-based cytology. Women with normal cytology are recalled for a repeat screen the following year, those with abnormal cytology are referred for colposcopy. Women recalled in year 2 follow the same pattern as year 1. Anyone HR-HPV positive in year 3 is referred for colposcopy [33]. The model included the cost of sample collection, HPV testing, cytology and colposcopy (where needed) for routine screening in year 1 and recall screening in years 2 and 3.

### Self-screening strategies

An opt-in rather than an opt-out strategy was assessed; individuals being invited to screen via letter with acceptance on an app or website (e.g., the NHS England app). Those who accept, receive a self-sampling kit in the mail containing the Colli-Pee® device [31] (Strategy 2) or the FLOQSwab® (COPAN Diagnostics Inc, Brescia, Italy) [34] (Strategy 3) plus instructions and a return Freepost envelope. Using the standard postal service, the sample is sent to a laboratory for HR-HPV testing. As in SoC, HR-HPV negative women are discharged to routine recall. Anyone HR-HPV positive is invited for cervical sampling so that cytology can be performed. This second sample is not tested for HPV. As in SoC, HR-HPV positive individuals with normal cytology are recalled for follow-up or referred for colposcopy (if in year 2). Recall testing in year 2/3 is via self-sampling followed by clinician-collected cervical sampling if HR-HPV positive.

### Outcomes

The primary outcomes assessed were the overall screening costs, the number of complete screens, and the average cost per complete screen (calculated as the total cost divided by the number of complete screens). A complete screen refers to either an HPV negative result or an HPV positive result with cytology and colposcopy or recall in years 2/3 where required.

The secondary outcomes assessed were the number of women screened, the number lost to follow-up (LTFU) and the cost per colposcopy (calculated as the total cost divided by the number of colposcopies). Each outcome was calculated for the complete 3-year screening cycle.

In scenario analyses, the total cost using each of the sampling strategies was calculated using the number of women invited to routine screening in the NHSCSP in 2020/21.

### Population

The same hypothetical cohort of 10,000 individuals was used for each sampling strategy. Data on HPV positivity, cytology, and colposcopy were taken from the NHSCSP [35] and a pilot study of 403,883 women screened in England [36,37]. Age-specific data from the study were adapted to reflect the national age distribution of people screened in the NHSCSP (Supplementary Tables 8-10). Since there are limited data to inform HPV positivity estimates in people who do not regularly screen, it was assumed that non-attenders had the same prevalence of HPV and abnormal cytology as attenders.

### Cost

Costs included: screening invite letter (including postage), sample collection in primary care, self-sample collection kit including instructions and return envelope, postage of self-collection kits and return of samples, laboratory HPV testing, cytology, and colposcopy (Table 1 and Supplementary Tables 1-5). Unit costs (presented in GBP £), were informed using published studies, NHS tariffs, Royal Mail postal charges, quotes from manufacturers, or estimated where necessary.

**Table 1.** Base case model input parameters and the high and low values used in the one-way deterministic sensitivity analysis (DSA).

| Parameter | Baseline value | Low value | High value | Reference/Comment |
| --- | --- | --- | --- | --- |
| **Cost data** |  |  |  |  |
| *All sampling strategies* |  |  |  |  |
| Invitation to screen | £0.42 | £0.34 | £0.50 | [39] |
| Cytology | £9.06 | £7.25 | £18.12 | [6,40] Supplementary Table 1 |
| Colposcopy<sup>1</sup> | £205.63 | £164.50 | £246.76 | [6,40] Supplementary Table 2 |
| Discount rate | 3.5% | 0% | 5% | [41] Standard rate recommended by NICE |
| *Clinician-collected cervical sampling* |  |  |  |  |
| Sample collection | £26.67 | £21.33 | £32.00 | [38,42,43] Supplementary Table 3 |
| HPV testing | £15.86 | £12.69 | £19.03 | [44] Supplementary Table 6 |
| *FV urine self-sampling* |  |  |  |  |
| Self-sample kit (including postage to individual) | £6.23 | £4.98 | £7.48 | Estimate provided by Novosanis<sup>2</sup>, Supplementary Table 4 |
| Return postage of sample to laboratory | £1.07 | £0.86 | £1.28 | [45] Supplementary Table 4 |
| HPV testing | £15.67 | £12.54 | £18.81 | [44] Supplementary Table 6 |
| *Vaginal self-sampling* |  |  |  |  |
| Self-sample kit (including postage to individual) | £3.61 | £2.89 | £4.33 | [4,46] Supplementary Table 5 |
| Return postage of sample to laboratory | £1.07 | £0.86 | £1.28 | [45] Supplementary Table 5 |
| HPV testing | £20.25 | £15.63 | £24.87 | [44] Supplementary Table 6 |
| **Probabilities (for women age 25 to 65 years)** |  |  |  |  |
| *All sampling strategies* |  |  |  |  |
| Uptake at invitation | 0.8191 | 0.6553 | 0.9829 | Calculated value<sup>3</sup> |
| HR-HPV positive result (Year 1) | 0.1155 | 0.0924 | 0.1386 | [6,36] Supplementary Table 9 |
| HR-HPV positive result (Year 2) | 0.5647 | 0.4518 | 0.6777 | [6,36] Supplementary Table 10 |
| HR-HPV positive result (Year 3) | 0.6578 | 0.5263 | 0.7894 | [6,36] Supplementary Table 10 |
| Cytology abnormal (Year 1) | 0.2759 | 0.2207 | 0.3311 | [6,36] Supplementary Tables 11 and 12 |
| Cytology abnormal (Year 2) | 0.1866 | 0.1492 | 0.2239 | [6,36] Supplementary Tables 11 and 12 |
| LTFU at HPV recall (Year 2/3) | 0.4464 | 0.3571 | 0.5356 | NHSCSP [6] Average value for 2017 to 2020 |
| LTFU at referral to colposcopy | 0.2520 | 0.2268 | 0.2772 | NHSCSP [6] Average value for 2017 to 2020 |
| *Clinician-collected cervical sampling* |  |  |  |  |
| Does not attend booked appointment | 0.1210 | 0.0968 | 0.5046 | [47] Assumption based on 12% non-attendance at booked appointments in primary care |
| *FV urine self-sampling* |  |  |  |  |
| Does not return sample | 0.1210 | 0.0968 | 0.3554 | [48] Assumption<sup>4</sup> |
| Attends for recall cervical sample following positive HPV result (Year 1 or 2) | 0.9120 | 0.7931 | 1.000 | [4,49] |
| *Vaginal self-sampling* |  |  |  |  |
| Does not return sample | 0.1210 | 0.0968 | 0.3554 | [48] Assumption<sup>5</sup> |
| Attends for recall cervical sample following positive HPV result (Year 1 or 2) | 0.9120 | 0.7931 | 1.000 | [4,49] |

<sup>1</sup> Weighted by the proportion with and without biopsy (Supplementary Table 2)
<sup>2</sup> Cost is dependent on the number of devices purchased.
<sup>3</sup> Back calculated based on 72% uptake of screening in NHSCSP in 2017-20 (Supplementary Table 7) and 12% non-attendance in individuals who book primary care appointments [47].
<sup>4</sup> Based on 12% non-attendance at booked appointments in primary care.
FV, first-void; HR-HPV, high-risk human papillomavirus; LTFU, lost to follow-up.

Where required, costs were inflated to 2020/2021 prices using the NHS cost inflation index (NHSCII) [38]. A discount rate of 3.5% was applied to costs incurred in years 2/3 (Figure 1). The cost of HPV testing for vaginal swabs or urine samples was adapted from the cost of testing cervical samples used in a published economic evaluation [29] to account for changes in the staff time and consumables required (Supplementary Table 6). These calculations were informed by published HPV assay protocols and with input from a Lead Scientist at one of the laboratories providing HPV testing for the NHSCSP.

For the self-sampling strategies, the model included the cost of self-sampling kits requested and sent but not used in year 1 but did not include the cost of unused kits in years 2/3. For SoC, the cost of non-attendance at booked appointments in primary care (for any year) was not included. The cost of an app or website used to select self-sampling was not included in the model. Nor was the cost of a reminder letter for non-responders, costs related to training or changes to laboratory equipment or costs for the administration or coordination of the NHSCSP.

### Probability inputs

Data from an English pilot study were used to inform HPV positivity in years 1-3 [36], age-weighted to represent the national age distribution of individuals screening within the NHSCSP in 2020-2021 [6] (Supplementary Tables 8-10). The same probabilities for HPV positivity were used for each sampling strategy since there is growing evidence that assays optimised for use on FV urine or vaginal swabs are non-inferior to existing assays used on clinician-collected cervical samples [22– 25,50–52] and therefore, it was assumed that HPV assays for each sample type had equivalent performance.

The probability of loss to follow-up at colposcopy and at HPV recall were informed by NHSCSP data over the period 2017 to 2020 [6]. The chance of attending for clinician-collected sampling following a positive HPV result was the same for both self-sampling strategies and informed by published studies [4,49].

Screening uptake was based on NHSCSP 2017-2020 data (Supplementary Table 7) and the same for each sampling strategy. The probability of booking an appointment (Strategy 1) or requesting a self-sampling kit (Strategy 2 and 3) was back-calculated to account for a 12% non-attendance or non-return of samples [47].

### Deterministic sensitivity analysis

A one-way DSA was performed to assess the impact of varying each parameter on the cost per complete screen. High and low values for costs and probabilities were informed by published studies or varied by +/-10% or +/-20% the baseline value. Due to uncertainty around the cost of cytology, +100% the baseline value was used for the high value.

### Scenario analysis

In scenario analyses, the total cost of the NHSCSP was calculated for each sampling strategy using a population of 4,039,982 representing the number of people invited to routine screening in 2020/21 [6]. Costs were also calculated for scenarios where the choice of self-sampling was offered to everyone or only to non-attenders and where the uptake was held constant or increased, combined with different levels of conversion to self-sampling, informed by studies predominantly focused on self-sampling offered to non-attenders (Supplementary Table 13).

Additional scenarios were used to assess the impact of lower HPV positivity i.e., in an HPV vaccinated cohort and higher HPV positivity as might be anticipated in some non-attender groups.

### Patient and public involvement

‘Non-invasive cervical screening’ was identified as an important unmet research need according to patients and clinicians in a recent James Lind Alliance Detecting Cancer Early Priority Setting Partnership [53]. A survey of more than 2,000 women found that 80% prefer non-invasive screening, rising to 88% amongst current non-attenders [54]. Whilst urine self-sampling may offer a ‘major breakthrough’ to ‘end smear fear’ [55], a formal cost evaluation is required to inform its implementation. Results from this study will be disseminated via social media and Jo’s Cervical Cancer Trust website [56].

## Results

### Base case results

The primary and secondary outcomes of the three sampling strategies are presented in Table 2. For the cohort of 10,000 women offered screening, the cost per complete screen was £56.81 for SoC, compared to £38.57 for FV urine sampling and £40.37 for vaginal self-sampling. Compared to SoC, this results in a cost-saving of £18.24 (32%) per completed screen for FV urine and £16.44 (28%) for vaginal self-sampling.

**Table 2.** Baseline results for comparison of three sampling strategies for cervical cancer screening.

| Outcomes for a cohort of 10,000 women age 25 to 64 over a 3-year screening cycle | Clinician-collected cervical sampling | FV urine self-sampling | Vaginal self-sampling |
| --- | --- | --- | --- |
| Number of complete screens | 6782 | 6738 | 6738 |
| Number attending for cervical sampling | 7618 | 915 | 915 |
| Number LTFU | 418 | 462 | 462 |
| Number who have a colposcopy | 240 | 213 | 213 |
| Total screening costs | £385,294 | £259,879 | £272,008 |
| Costs related to sampling<sup>1</sup> | £207,067 | £89,970 | £67,528 |
| Costs related to laboratory testing<sup>2</sup> | £129,708 | £126,722 | £161,293 |
| Cost per complete screen<sup>3</sup> | £56.81 | £38.57 | £40.37 |
| Cost per colposcopy<sup>4</sup> | £1,608 | £1,220 | £1,277 |

<sup>1</sup>Includes costs for screening invitation, sample collection, self-sampling kits and kit postage.
<sup>2</sup>Includes costs for laboratory HPV testing and cytology.
<sup>3</sup>Calculated as the total cost of screening divided by the number of complete screens.
<sup>4</sup>Calculated as the total cost of screening divided by the number of colposcopies.
Complete screen refers to either an HPV negative result or an HPV positive result with cytology and colposcopy where required plus recall in year 2/3 where required.
FV, first-void; LTFU, lost to follow-up refers to those who receive a positive HPV result but do not attend for further investigations.

Since there are more steps in the self-sampling strategies (because HR-HPV positive women must return to provide a sample for cytology), more people were LTFU than in SoC (n=462 vs. n=418 respectively) and fewer women had a colposcopy. Using the base case values for attendance and LTFU, screening uptake would need to be 81% in the self-sampling strategies for the same number of women to have colposcopy in each strategy. At this uptake, the total screening costs for the cohort were £291,807 for FV urine sampling and £305,451 for vaginal sampling and the cost per complete screen was £38.50 for FV urine sampling and £40.30 for vaginal sampling.

### Sensitivity analysis

DSA was performed for each sampling strategy. Parameters with the most effect on the cost per complete screen are presented in Figure 2. For all three sampling strategies, the cost of HPV testing, the cost of colposcopy and the probability of HPV positivity had a large effect on the cost per complete screen. In SoC, the cost of sample collection had the greatest effect when altered and for the self-sampling strategies, the probability of not returning a sample was important.

**Figure 2.**
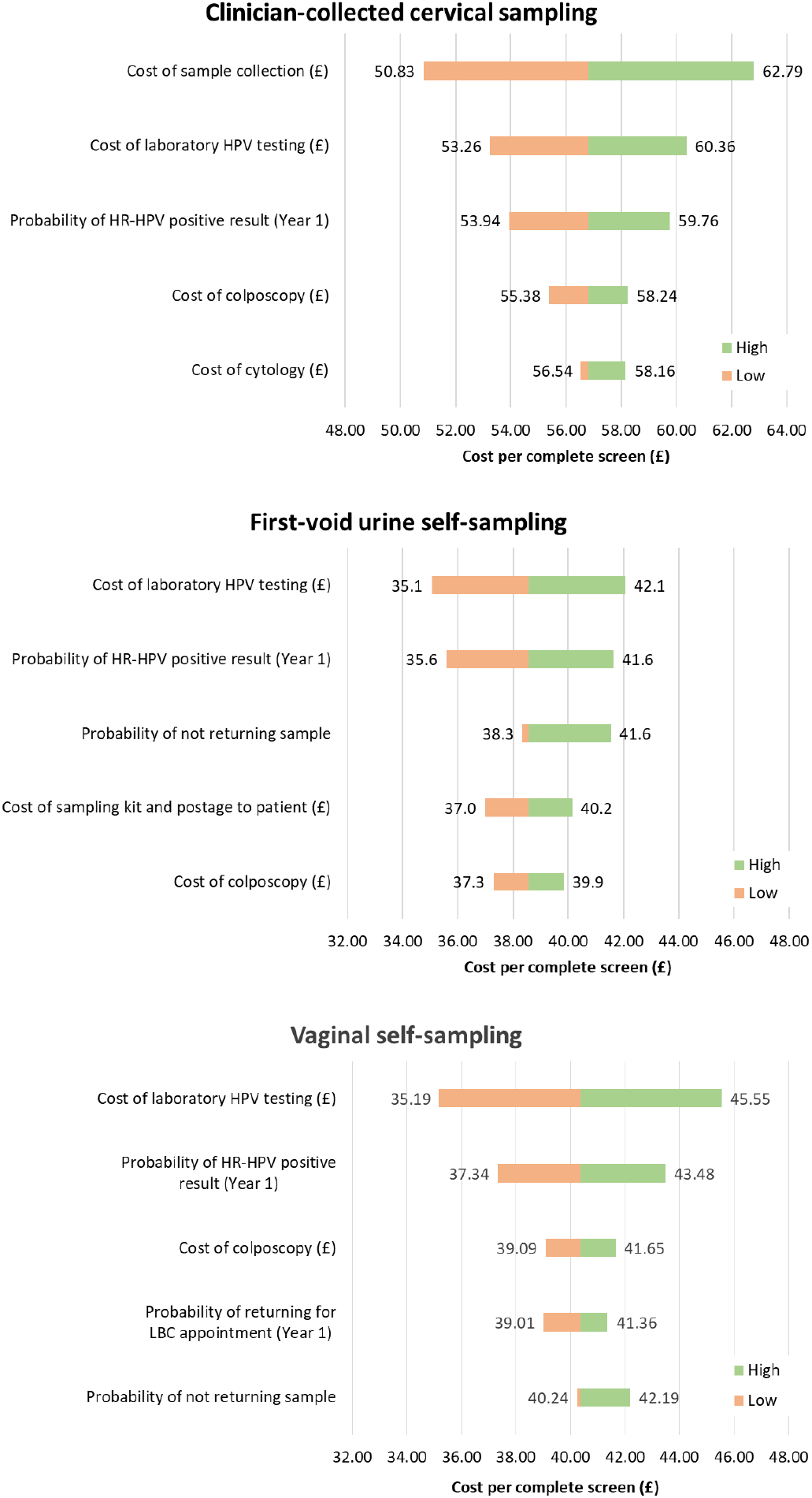
Tornado plots of univariate sensitivity analysis presenting the five parameters that led to the greatest change in the cost per complete screen (£) for each sampling strategy.

### Scenario analysis

Several scenarios were assessed (Table 3 and Supplementary Tables 13-14). Using the number of people invited to routine screening in 2020/21, [6] the total cost for each sampling strategy (assuming the same uptake, Scenario 1) for 4,039,982 people offered screening would be £155,658,155 for SoC, £104,990,567 for FV urine sampling, and £109,890,829 for vaginal self-sampling.

**Table 3.**
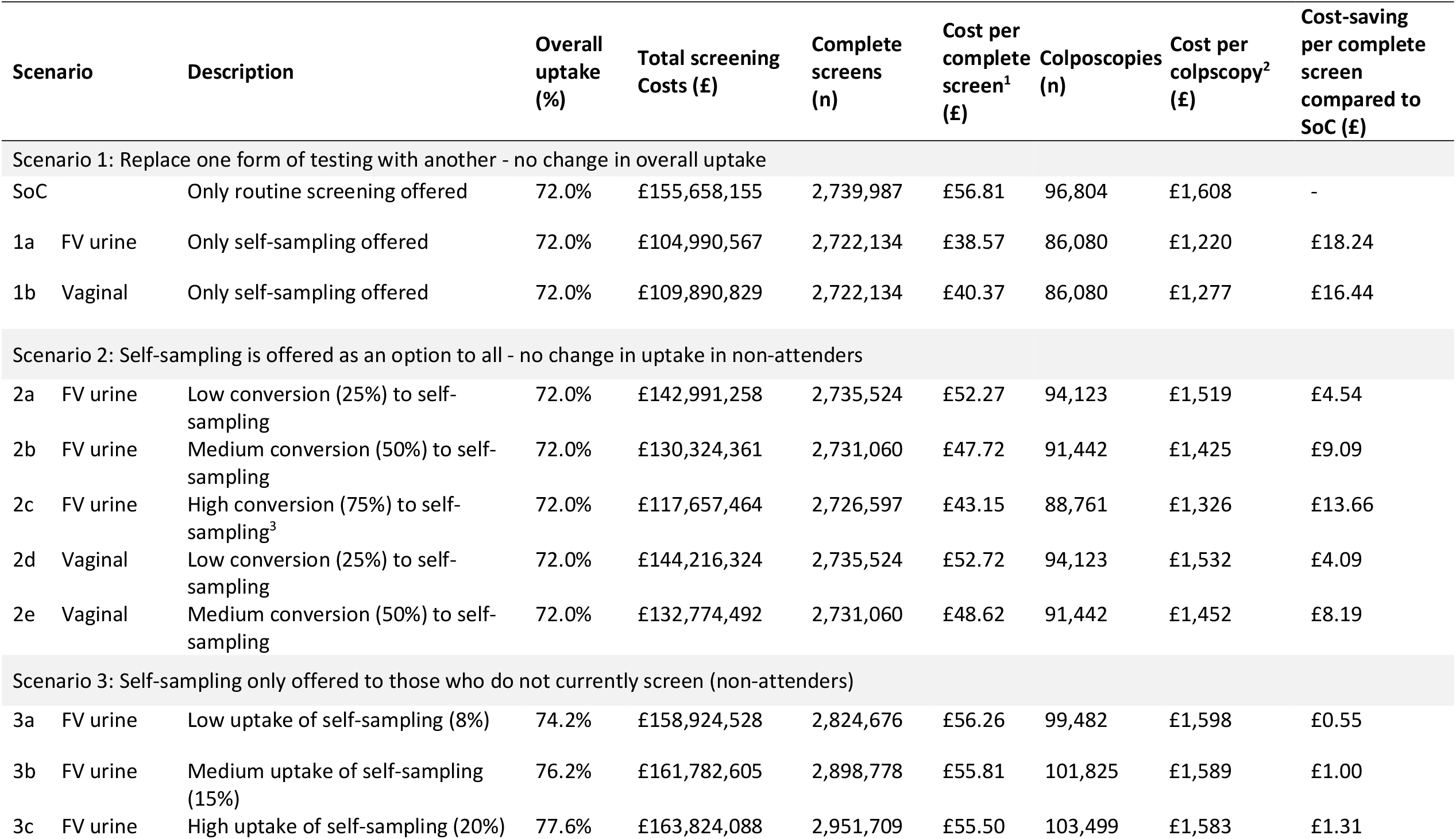
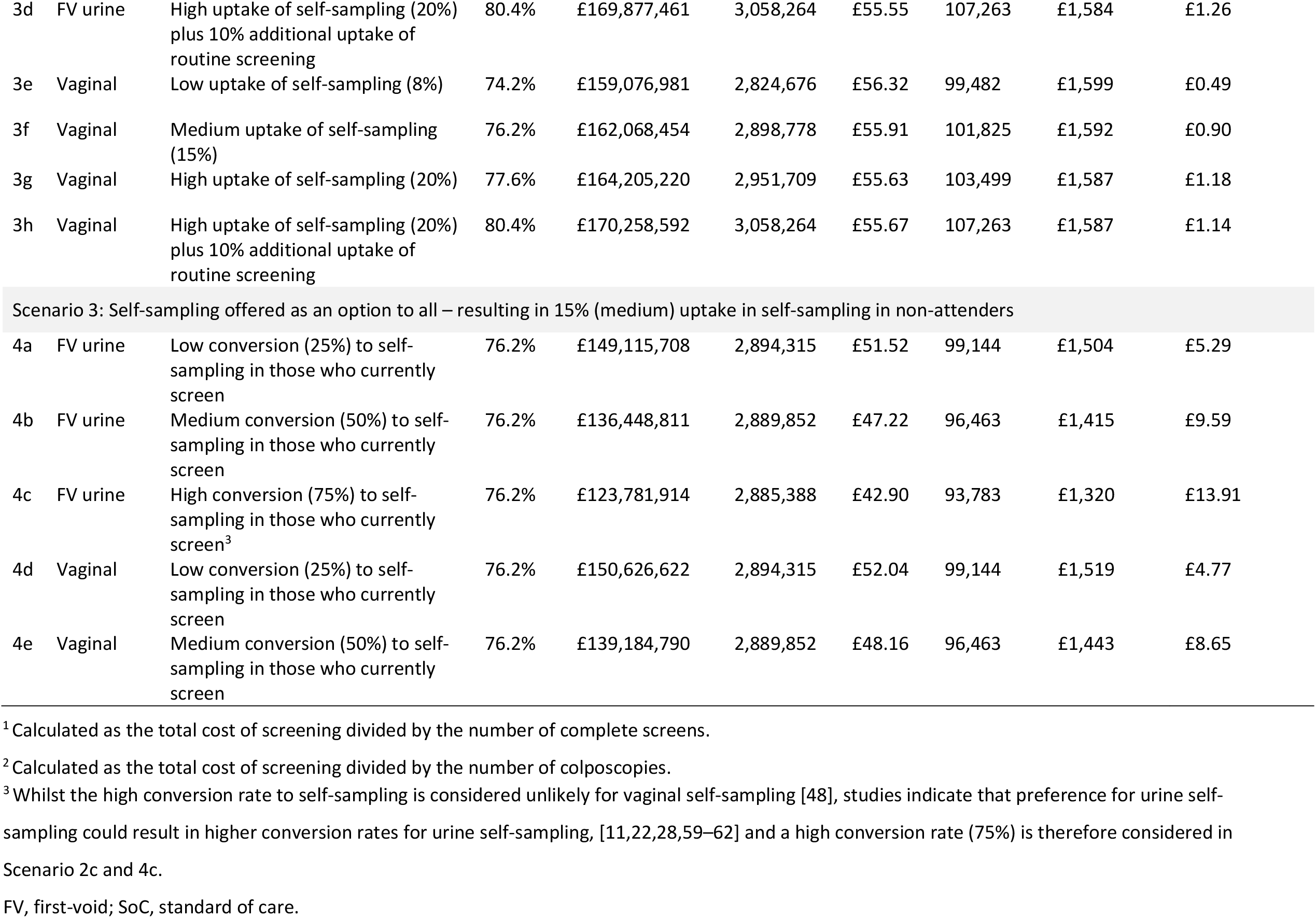
Overall screening costs for England under different screening scenarios for 4,039,982 people offered screening.

Offering self-sampling to non-attenders (Scenario 3) would result in a cost of £56.26 per complete screen for FV urine and £56.32 for vaginal self-sampling (with 8% uptake), less than in SoC (£56.81). Although the cost per test was less for self-sampling than for SoC, the overall cost of the NHSCSP would increase if offering self-sampling meant that the overall uptake increased, and the amount of clinician-collected sampling was unchanged (Scenario 3). If there was no change in uptake but some people opted for self-sampling instead of clinician-collected sampling, then the overall cost would fall but, due to the additional steps in the self-sampling pathway (in our current model), the total number with a complete screen would also fall (Scenario 2). Scenario 4 had a more favourable outcome where the offer of self-sampling resulting in 15% uptake in non-attenders plus some conversion to self-sampling in attenders. With 50% conversion (Scenario 4a, informed by a recent preference study in England [57]), the overall number receiving a complete screen would be higher than in SoC and save £19.2 million (FV urine) or £16.5 million (vaginal self-sampling) over the 3-year screening cycle for this cohort of 4-million women.

In England, HPV vaccination has been offered to girls of age 12 to 13 since September 2008. As such, the first cohort of vaccinated girls is now eligible for the NHSCSP [58]. A recent study found a 23% decrease in HPV positivity in 2018 in a cohort in which 55% had received three doses of the bivalent HPV vaccine, compared to a same-aged cohort in 2013 with 0% vaccinated [58]. If HPV positivity was 23.1% lower at baseline and the year 2 and year 3 recall (Supplementary Table 15) the cost per complete screen would be £52.72 for SoC, £34.43 for FV urine sampling, and £36.17 for vaginal self-sampling (Supplementary Table 16).

A higher HPV prevalence might be expected in some non-attenders. The cost per complete screen for the FV urine self-sampling (£42.74) and vaginal self-sampling (£44.59) was less than the cost for SoC (£60.99) even when the high HPV positivity values (used in the DSA) were used in years 1, 2 and 3.

## Discussion

### Main findings

The results of the base case model indicate that self-sampling for cervical cancer screening costs less than clinician-collected sampling assuming equivalent performance of HPV testing on self-collected and clinician-collected samples. The magnitude of savings to a screening programme is dependent on whether self-sampling is offered to non-attenders or to everyone eligible for screening, and the impact on uptake. The self-sampling pathway currently requires an additional step for clinician-collected sampling following a positive HPV result which might be avoidable with newer technologies such as DNA methylation. This additional step means that overall uptake must be higher to result in the same number of complete screens as SoC due to LTFU.

In some of the scenarios assessed, the NHSCSP would make considerable savings whilst increasing the number of complete screens. With the HPV vaccinated cohort entering the NHSCSP, the HPV positivity is anticipated to fall and would further reduce the costs, particularly in self-sampling pathways where a negative HR-HPV result does not require an appointment.

### Strengths and limitations

This is the first study to model the costs of vaginal swab and FV urine self-sampling within the NHSCSP. As such it provides valuable data to inform decision making about the future use of self-sampling in England, elsewhere in the UK, and other countries with similar screening programmes.

As with all models, several assumptions were made for simplicity or due to the sparsity of data available to inform the parameters. One crucial assumption is that HPV testing of self-collected samples has (or will have) equivalent performance to clinician-collected samples. This assumption was made based on previous evaluations indicating non-inferiority of sensitivity for CIN2+ on self-collected samples when using PCR-based assays [22,24,50–52]. Further optimisation of HPV assays for self-collected samples is anticipated if self-sampling were incorporated into screening programmes. There are a number of ongoing trials that will provide real-world data supporting this assumption [63–66]. If the sensitivity for CIN2+ detection is inferior for self-collected samples, more cases would be missed than in SoC and therefore it would be most appropriate to offer self-sampling to non-attenders who would otherwise not screen.

In a high uptake setting such as England, there is a risk in changing well-established screening programmes since shifting to self-sampling could result in a temporary or long-term drop in uptake. It is not sufficient for HPV tests to demonstrate high performance it is crucial that people offered screening are informed about and have confidence that testing is highly accurate to encourage uptake and confidence in the screening programme. It will be important to collect real-world evidence, provide education/awareness programmes for women and clinicians and evaluate changes carefully to ensure that overall uptake is maintained or improved to further improve the detection and treatment of cervical cancer and CIN. In the context of increasingly vaccinated cohorts entering the NHSCSP with lower HPV prevalence, the cost and cost-effectiveness of the programme will decrease and therefore solutions such as self-sampling (and potentially DNA methylation triage on the same sample in place of cytology) will be of great interest to programme managers and decision-makers as they can reduce the costs of the programme and the burden on healthcare services.

Where costs for England were calculated (in scenario analyses) it was assumed that HPV positivity was the same in attenders and non-attenders. This is unlikely to be the case; studies indicate a higher prevalence of CIN2+ and cervical cancer in long-term non-screening groups compared to populations who regularly screen [67–69]. However, the results of the DSA (Figure 2) and scenario analysis indicate that self-sampling would cost less than clinician-collected sampling even in a population with higher HPV positivity.

In our base case model, fewer people received colposcopy in the self-sampling strategies due to the additional LTFU at the point where HPV positive people must attend to provide a sample for cytology. There were limited data on the probability of returning self-samples and attendance for cervical sampling following an HPV positive result [68], both of which will be informed by future population-based evaluations and may change over time if self-sampling were widely promoted and became ‘the norm’. Variations in these probabilities impact the cost per screen but self-sampling remains the cheaper option even when these probabilities were low (Figure 2). Future developments enabling molecular triage of self-collected samples for colposcopy referral would simplify the pathway and eliminate this risk of LTFU [63,70]. Its impact on cost per screen is unclear as it would largely depend on the cost of molecular triage, which is not currently approved for use within the NHSCSP.

### Interpretation

Previous cost evaluations of self-sampling for HPV testing support our finding that self-sampling costs less than routine cervical screening [27]. As a strategy to offer to non-attenders who decline routine screening, self-sampling may increase national screening coverage closer to the target of 80% [71] reversing the downward trend in screening uptake seen over the past decade and during the COVID-19 pandemic. Introduced as a choice for all, self-sampling has the potential to save money and increase coverage.

If self-collected samples require even a few more minutes of laboratory staff time per sample, this may be impractical when scaled up to 2500 samples a day, the current throughput at one of the NHSCSP laboratories [personal communication]. Therefore, full rollout to everyone eligible for screening is dependent on the extent to which laboratory procedures can be automated for testing self-collected samples [72].

There may be hesitancy in introducing a change to screening for the whole eligible screening population. A 2017 Dutch study assessing the switch from cytology to primary HR-HPV screening with the simultaneous introduction of vaginal self-sampling as an option offered to all observed a decrease in participation (from 64% to 61%) [73]. Other recent changes to the delivery of healthcare and the widespread use of self-testing in the context of the COVID-19 pandemic may play a role in increasing the acceptability of self-sampling. Furthermore, there is evidence that urine self-sampling is more acceptable than vaginal self-sampling due to it being less invasive and easier to collect [74] and women feeling more confident providing a urine sample compared to a vaginal sample [75]. As such, self-sampling using urine may align with patient preferences, address screening inequalities and remove some barriers to clinician-collected cervical and/or vaginal self-sampling [11,22,28,59– 62].

### Future research

Our model represents one pathway for self-screening. There are alternative pathways, self-sampling devices, and ways to distribute self-sampling kits, some of which have been assessed in trials. For example, self-sampling could be offered to non-attenders after an initial offer of routine screening is ignored [76] either by sending another invite or the self-collection kit [77]. Our model used an opt-in strategy for the initial screen and an opt-out approach for the 12-month recall. One alternative is an opt-out strategy for the initial invite (i.e., sending the self-sample kit with the invite), a strategy successfully used in the UK for bowel cancer screening [14]. Opt-out strategies have the advantage of increasing uptake but increase the initial costs due to the wastage of unused kits [51]. Alternatives to a letter invitation may also be modelled, for example, offering self-sampling within primary care or community settings. An Australian study found high uptake (86%) in women who were offered vaginal self-sampling for HPV screening while attending a GP appointment [78]. Uptake was 68% (of which 66% returned the sample) in a study conducted in England where women could either collect the self-sample while at the clinic or after returning home [79]. Real-world evaluations to assess the best way to offer screening would provide invaluable insight, particularly if mixed methods were used to understand the reasons for any change in uptake. An evaluation of whether an HPV test on the second sample taken for cytology is of benefit would also inform this part of the screening pathway if self-sampling were adopted.

In our model, for each sampling strategy, the same type of sampling was used for routine and recall screening. Alternatively, clinician-collected sampling could be used (for all strategies) in years 2 and 3, since HPV prevalence is considerably higher in the recall group.

The increase in recent and ongoing research activities around urine self-sampling for HPV testing [64,65,80–83] indicates that there is a lot of interest in its potential as an alternative method of cervical screening. To further inform screening programmes, further research is needed to optimise urine collection, the volume of urine collected, transport media, and compatibility of new or existing HPV assays. The screening programme will need to evolve as the cohort changes with increasing numbers vaccinated against HPV and a reduction of HPV infection, pre-cancerous lesions, and cervical cancer.

## Conclusion

Cost is not a barrier to the use of self-sampling within the NHSCSP. Self-sampling for primary HPV testing should be considered as it provides a less costly alternative to routine clinician-collected cervical screening with the additional benefit of improving user experience and choice.

Many countries are considering how best to implement self-sampling options for HPV primary testing. This study provides evidence on the scale of cost savings which could be realised and directly compares two self-sampling methods.

## Data Availability

All data relevant to the study are included in the article or uploaded as supplementary material. The model is not publicly available.

## Author statement

## Acknowledgements

We would like to thank and acknowledge the contribution of Suzanne Carter at Manchester University NHS Foundation Trust. Additionally, we would like to thank Dr Mathilde Vankelegom and Anne Meiwald for reviewing the model structure and input data used in the analysis.

## Disclosure of interest

SH, KPS, VS, KT & EA are employed by Aquarius Population Health which received funding for this study. Aquarius Population Health works on projects related to diagnostics for different commercial and academic clients and as part of grant-funded projects. EJC is a National Institute for Health Research (NIHR) Advanced Fellow (NIHR300650), and her work is funded by the NIHR Manchester Biomedical Research Centre (IS-BRC-1215-20007). AS is a Clinical Scientist employed by the NHS working within the NHSCSP in the capacity of HPV Lead Scientist and Pathway Manager.

## Contribution to authorship

SH, KPS, and VS designed the study and model structure with support and input from KT and EJC. KPS built the main model and ran the analyses. VS developed the scenarios analysed with input from SH. KPS, VS, and SH identified data to inform the model parameters and transformed these where necessary. AS helped inform model parameters. EJC led on patient and public engagement. SH, KPS, VS, KT and EJC contributed to data interpretation. The paper was drafted by SH, KPS, VS, and EJC with input from AS, KT and EA. The project was overseen by SH, with input from KT and EA.

## Details of ethics approval

No ethical approval was required or sought as only secondary data sources were used, there was no randomisation or change to patient care, and no patient identifying information was obtained or used.

## Funding

This study was funded by Novosanis, a subsidiary of OraSure Technologies Incorporated. The design, results, and interpretation of the study were generated independently by the authors. Novosanis was given the opportunity to review the manuscript for medical and scientific accuracy.

