## Supplementary material for "An economic evaluation of two self-sampling strategies for HPV primary cervical cancer screening compared with clinician-collected sampling"

### Contents

Section 1: Calculations used to inform unit costs

Supplementary Table 1. Variables used to inform the cost of cytology

Supplementary Table 2. Variables used to inform the cost of colposcopy

Supplementary Table 3. Variables used to inform the cost of sample collection: clinician-collected cervical smear

Supplementary Table 4. Variables used to inform the cost of FV urine self-sample collection

Supplementary Table 5. Variables used to inform the cost of sample collection for each sampling strategy

Supplementary Table 6. Variables used to inform the cost of HPV testing for each sampling strategy

Section 2: Calculations used to inform probabilities

Supplementary Table 7. Variables used to inform the uptake of cervical screening

Supplementary Table 8. Number of women tested for HPV in England 2020/2021 by age category

Supplementary Table 9. Raw and weighted baseline HPV positivity

Supplementary Table 10. Raw and weighted HPV positivity at year 2 (12-month recall) and year 3 (24-month recall)

Supplementary Table 11. Variables used to calculate the percentage of screens resulting in abnormal cytology result

Supplementary Table 12. Variables used to inform the probability of receiving an abnormal cytology diagnosis if HR HPV positive

Section 3: Calculating the overall screening costs for England given self-screening offered to different groups and a range of plausible uptake rates

Supplementary Table 13. Scenarios used to calculate overall screening costs

Supplementary Table 14. Break-down of calculations in scenarios

Section 4: Calculating cost per complete screen given low and high values for HR HPV positivity

Supplementary Table 15. Variables and calculations used to inform the low values in HPV infection in scenario analysis

Supplementary Table 16. Cost per complete screen - results from the scenario analysis using low values for HR HPV infection

**Section 1: Calculations used to inform unit costs**

**Supplementary Table 1. Variables used to inform the cost of cytology**

| Variable | Value | Reference/Comment |
|---|---|---|
| Cost of cytology testing by result[1] | | |
| Negative result | £6.74 | ARTISTIC trial, 2014 (Table 88) [1] |
| Low-grade dyskaryosis | £19.03 | ARTISTIC trial, 2014 (Table 88) [1] |
| High-grade dyskaryosis | £19.23 | ARTISTIC trial, 2014 (Table 88) [1] |
| Percentage of patients with each cytology test result | | |
| Negative result | 81.2% | English pilot study Table 1 [2]; NHSCSP Table 6. [3] Calculated by adjusting the proportion of different cytology results in the NHSCSP data tables by HPV positivity data at baseline from the English pilot study [2]. |
| Low-grade dyskaryosis | 14.2% | |
| High-grade dyskaryosis | 4.6% | |
| **Weighted cost** | **£9.06** | Calculated by weighting the cost of cytology by the proportion of patients with each cytology result. |

[1] Costs inflated from 2009/10 to 2020/2021 value.

**Supplementary Table 2. Variables used to inform the cost of colposcopy**

| Variable | Value | Reference/Comment |
|---|---|---|
| Colposcopy without biopsy | £199.00 | NHS Tariff 2021/22 for diagnostic colposcopy outpatient procedure (HRG code MA38Z) [4] |
| Colposcopy with biopsy | £214.00 | NHS Tariff 2021/22 for diagnostic colposcopy with biopsy outpatient procedure (HRG code MA39Z) [4] |
| Percentage of patients referred to colposcopy who receive a biopsy | 44.2% | NHSCSP 2021/22, Table 23 [3] |
| **Weighted cost** | **£205.63** | Calculated by weighting the cost of colposcopy by the proportion of patients who receive a biopsy. |

**Supplementary Table 3. Variables used to inform the cost of sample collection: clinician-collected cervical sampling**

| Variable | Value[1] | Reference/Comment |
|---|---|---|
| Staff costs | | |
| Staff consultation time | 13 minutes, 45 seconds | LBC/HPV pilot study, 2004 (Table 4A.2) [5] |
| Nurse cost per hour | £44.00 | PSSRU, 2020/21 estimate for nurse cost per hour [6] |
| GP cost per hour | £255.00 | PSSRU, 2020/21 estimate for GP cost per hour of patient contact [6] |
| Percentage of cervical samples collected by a nurse | 80% | LBC/HPV pilot study, 2004 [5] |
| Percentage of cervical samples collected by a GP | 20% | LBC/HPV pilot study, 2004 [5] |
| Weighted cost per minute | £1.44 | Weighted by the proportion of samples taken by each type of staff. |
| Total staff costs | £19.75 | |
| Sample transport to laboratory | £1.31 | ARTISTIC trial, 2014 (Table 89) [1] |
| ThinPrep consumables pack | £5.60 | LBC/HPV pilot study, 2004 (Table 4A.6) [5] |
| **Total cost** | **£26.67** | |

[1] Costs inflated to 2020/2021 value where required. GP, general practitioner.

**Supplementary Table 4. Variables used to inform the cost of FV urine self-sample collection**

| Variable | Value[1] | Reference/Comment |
|---|---|---|
| FV urine self-sampling | | |
| Colli-Pee® device (10mL with UCM™) | £2.41 | Estimate provided by Novosanis. Cost varies according to quantity purchased. |
| Self-collection kit (includes return cardboard envelope, safety bag containing absorbent tissue, and user instruction pamphlet). | £1.40 | Estimate provided by Novosanis. Cost varies according to quantity purchased. |
| Postage of self-collection kit to individual | £2.42 | Based on 2nd class postage using Royal Mail 48® (Small parcel up to 1kg). [7] |
| **Total kit cost** | **£6.23** | |
| Return postage of sample to laboratory | £1.07 | Based on 1st class return postage (large letter up to 100 grams). [8] Included in model with HPV assay cost i.e., only for those who return a sample. |

[1] Costs inflated to 2020/2021 value where required.

**Supplementary Table 5. Variables used to inform the cost of vaginal swab self-sample collection**

| Variable | Value[1] | Reference/Comment |
|---|---|---|
| Vaginal swab self-sample | | |
| COPAN FLOQSwab® | £1.38 | Estimate based on price of Evalyn Brush vaginal self-sampling device (STRATEGIC trial, 2016) and wholesale prices for the COPAN FLOQSwab available online. [9–13] |
| Self-collection kit (includes return cardboard envelope, safety bag containing absorbent tissue, and user instruction pamphlet). | £1.40 | Estimate provided by Novosanis so self-collection kit used with the Colli-Pee® device[2]. |
| Postage of self-collection kit to individual | £0.83 | Based on 2nd class postage using Royal Mail 48® (large letter). [7] |
| **Total kit cost** | **£3.61** | |
| Return postage of sample | £1.07 | Based on 1st class return postage (large letter up to 100 grams). Included in model with HPV assay cost i.e. only for those who return a sample. [8] |

[1] Costs inflated to 2020/2021 value where required.

[2] Assumed that contents and pricing of a vaginal self-collection kit would be the same as the urine self-collection kit.

**Supplementary Table 6. Variables used to inform the cost of HPV testing for each sampling strategy**

| Variable | Value[1] | Reference/Comment |
|---|---|---|
| Clinician-collected cervical sample | | |
| HPV test | £15.86 | Legood, 2012 (Table 4) [14] |
| First-void urine sample | | |
| HPV test | £15.86 | Legood, 2012 (Table 4) [14] |
| Band 3 staff time saved[2] | 22 seconds | Expert communication. |
| Cost of staff time saved | £0.19 | PSSRU 2020/21 [6], based on staff cost per hour of £29.87 estimated for Band 3 scientific and professional staff calculated by subtracting ratio of pay difference (cost per working hour) between Band 4 and 5 staff from Band 4 cost per working hour. |
| **Total cost** | **£15.68** | |
| Vaginal swab sample | | |
| HPV test | £15.86 | Legood, 2012 (Table 4) [14] |
| Additional Band 3 staff time[3] | 5 minutes | Expert communication. |
| Cost of additional staff time | £2.49 | See comment for FV urine sample |
| Additional consumables[4] | £1.90 | Estimated cost for 20mL of PreservCyt liquid solution from UNICEF Supply Division. [15] |
| **Total cost** | **£20.25** | |

[1] All costs inflated to 2020/2021 value. Values indicated are per sample.

[2] Assumed that high-throughput testing platforms are compatible with the tubes used for self-collection of urine samples using the Colli-Pee® device and tubes can be placed directly into the racks placed in the machines thereby avoiding the pre-analysis stage. The pre-analysis stage involves loading 80

samples onto the pre-analysis machine (20 minutes) and unloading 80 samples (10 minutes) which equates to a total of 22.5 seconds per sample and a cost saving of 18.7p for staff time.

[3] Assumed that laboratory processing of self-collected vaginal samples using the COPAN FLOQSwab® dry swabs require additional time compared with clinician-collected cervical samples as specimens must be resuspended in preservative liquid solution before HPV assay analysis.

[4] Assumed that 20mL of PreservCyt liquid solution will be used as a buffer solution for resuspension.

**Section 2: Calculations used to inform probabilities**

**Supplementary Table 7. Variables used to inform the uptake of cervical screening**

| Year | 2017 | 2018 | 2019 | 2020 | Mean for 2017 to 2022 | Reference/Comment |
|---|---|---|---|---|---|---|
| Cervical screening uptake (%) | 72.0% | 71.4% | 71.9% | 72.2% | 71.9% | NHSCSP, Table 1. [16] Coverage is calculated as the proportion of eligible women (aged 25-64) screened within 3.5/5.5 years. |
| Number invited to early repeat recall (surveillance) | 388,374 | 317,953 | 283,049 | 310,389 | - | NHSCSP, Table 4. [16] Surveillance invitation of eligible individuals (aged 25-64) includes rescreen due to a previous positive HR HPV test and negative cytology. |
| Number attending early repeat recall (surveillance) | 215,982 | 177,818 | 161,851 | 163,694 | - | NHSCSP, Table 5. [16] Surveillance invitation of eligible individuals (aged 25-64) includes rescreen due to a previous positive HR HPV test and negative cytology. |
| Uptake of early repeat recall (surveillance) | 55.6% | 55.9% | 57.2% | 52.7% | 55.4% | NHSCSP, Table 4 and 5. [16] Calculated by dividing the number who attended by the number invited in the surveillance group. |
| Uptake of colposcopy | 75.6% | 74.6% | 74.4% | 74.6% | 74.8% | NHSCSP, Table 22. [16] Mean attended colposcopy appointments, following positive HPV and cytology testing. |

**Calculating the probability of an HR-HPV positive result in years 1, 2 and 3**

The English pilot study reported the proportion of women who were HR-HPV positive according to the age categories 24-29, 30-39, 40-49, and 50-64 years (Supplementary Table 9). The overall HPV positivity at baseline and at follow-up were calculated using data from the NHSCSP to inform the national proportion of people testing in England within these age groups (Supplementary Table 8).

**Supplementary Table 8. Number of women tested for HPV in England 2020/2021 by age category**

| Age group | Women HPV tested following routine screening invite (Year 1) | | Women HPV tested following recall (surveillance) screening invite (Year 2/3) | | Reference/Comment |
|---|---|---|---|---|---|
| | n | % | n | % | |
| 25-29 years | 346,695 | 15.6% | 36,747 | 17.9% | NHSCSP Table 5 [3] |
| 30-39 years | 661,483 | 29.9% | 84,011 | 40.9% | |
| 40-49 years | 633,036 | 28.6% | 48,208 | 23.5% | |
| 50-64 years | 574,786 | 25.9% | 36,249 | 17.7% | |
| All age groups combined | 2,216,000 | - | 205,215 | - | |

**Supplementary Table 9. Raw and weighted baseline HPV positivity**

| Age group | Proportion of women HR-HPV positive | NHSCSP age weighting | Reference/Comment |
|---|---|---|---|
| 25-29 years | 26.9% | 15.6% | English pilot study [2] |
| 30-39 years | 13.0% | 29.9% | |
| 40-49 years | 7.30% | 28.6% | |
| 50-64 years | 5.30% | 25.9% | |
| **Baseline HPV positivity for women 25 to 64 years** | **11.5%** | | Calculated using NHS age weighting from Supplementary Table 8 |

The English pilot study also reported the proportion of women with a persistent HR-HPV infection at 12- and 14-month follow-up. These data were used to inform the probability of an HR HPV positive result at the year 2 and year 3 rescreen (Supplementary Table 10) (i.e., at 12-month and 24-month follow-up after an HPV positive, cytology negative result in the previous year) using the national age weighting for women invited to early repeat recall (Supplementary Table 8).

**Supplementary Table 10. Raw and weighted HPV positivity at year 2 (12-month recall) and year 3 (24-month recall)**

| Age group | Proportion of women HR-HPV positive at year 2 | NHSCSP age weighting | Proportion of women HR-HPV positive at year 3 | NHSCSP age weighting | Reference/Comment |
|---|---|---|---|---|---|
| 25-29 years | 61.4% | 17.9% | 64.2% | 17.9% | English pilot study [2] |
| 30-39 years | 56.2% | 40.9% | 64.2% | 40.9% | |
| 40-49 years | 51.2% | 23.5% | 65.3% | 23.5% | |
| 50-64 years | 59.1% | 17.7% | 71.7% | 17.7% | |
| **HPV positivity 25 to 64 years** | **56.5%** | | **65.8%** | | Calculated using NHS age weighting from Supplementary Table 8 |

**Calculating the probability of different cytology results**

Data on outcome of reflex cytology following HR HPV testing were extracted from the NHSCSP published data. [3] The NHSCSP tables do not distinguish between HR HPV and cytology outcomes. This means the group reported as 'Negative' consists of (predominantly) people who were HR HPV negative, and did not receive reflex cytology, and a smaller number of people who were HR HPV positive with a 'normal' (i.e., negative) cytology result (Supplementary Table 11). Data from pilot study in England [2] were used to calculate the combined probability of being cytology abnormal if HR HPV positive (Supplementary Table 12).

**Supplementary Table 11. Variables used to calculate the percentage of screens resulting in abnormal cytology result**

| Screening result | | Routine screen | | Rescreen | Reference/Comment |
|---|---|---|---|---|---|
| | | First invite | Routine repeat invite | Surveillance | |
| Inadequate screen | | 2,364 | 10,701 | 2,477 | Data from the NHSCSP Table 6. [3] |
| Negative[1] | a | 239,826 | 1,892,906 | 181,377 | |
| Borderline changes | b | 4,306 | 14,427 | 7,138 | |
| Low-grade dyskaryosis | c | 10,810 | 24,038 | 9,612 | |
| High-grade dyskaryosis (moderate) | d | 2,023 | 5,996 | 2,406 | |
| High-grade dyskaryosis (severe) | e | 1,973 | 5,801 | 1,987 | |
| High-grade dyskaryosis (possible carcinoma) | f | 116 | 143 | 27 | |
| Possible glandular neoplasia (endocervical) | g | 63 | 507 | 189 | |
| Total number of people screened (a-g) | x | 259,117 | 1,943,818 | 202,736 | Excludes inadequate screens |
| Total number with abnormal cytology (b-g) | y | 19,291 | 50,912 | 21,359 | |
| Total number with high-grade cytology (d-g) | z | 4,175 | 12,447 | 4,609 | |
| Percentage of screens HR-HPV positive and abnormal cytology (y/x) | | 7.4% | 2.6% | 10.5% | Used in Supplementary Table 12 |
| | | 3.2% (combined) | | | |
| Percentage of abnormal cytology results that are high-grade (z/y) | | 21.6% | 24.5% | - | Used to inform cost calculations |
| | | 23.7% (combined) | | | |

[1]Assumption that this includes (predominantly) HR-HPV negative results as well as a small number of HR-HPV positive and cytology ‘normal’ results.

**Supplementary Table 12. Variables used to inform the probability of receiving an abnormal cytology diagnosis if HR HPV positive**

| Screening outcome | | Percentage | Reference/Comment |
|---|---|---|---|
| Routine screening | | | |
| Percentage of screens HR-HPV positive | a | 11.5% | English pilot study [2] Positive HPV result in year 1/baseline, calculated in Supplementary Table 9. |
| Percentage of screens HR-HPV positive and abnormal cytology | b | 3.2% | Calculated in Supplementary Table 11. |
| Percentage with abnormal cytology diagnosis among HR HPV positive screens | | 27.6% | Calculated variable (b/a). |
| Rescreen | | | |
| Percentage HR-HPV positive | c | 56.5% | English pilot study [2] Positive HPV result at 12 month recall, calculated in Supplementary Table 10. |
| Percentage with abnormal cytology | d | 10.5% | Calculated in Supplementary Table 11. |
| Percentage with abnormal cytology diagnosis among HR HPV positive screens | | 18.7% | Calculated variable (d/c). |

**Section 3: Calculating overall costs for a screening programme given offer of self-screening to different groups and a range of uptake rates**

The total cost of screening in England given different screening scenarios was assessed (Supplementary Table 14) using a total population of 4,039,982 individuals eligible for screening based on the number invited for routine screening in the NHSCSP in 2020/21 [3].

As well as the total costs, a breakdown of costs was presented for individuals adequately screened (i.e., not lost to follow-up [LTFU]), individuals not screened and individuals with incomplete screening (i.e., LTFU either because they did not attend for a cervical sample after a positive HPV result on their self-collected sample or for colposcopy or recall in year 2/3) (Results presented in the Main paper).

In Scenario 1, the current use of clinician-collected cervical samples would be completely replaced by one form of self-sampling, either FV urine (Scenario 1a) or vaginal swab (Scenario 1b).

In Scenarios 2 and 4, a choice is given to have routine screening or to self-sample. Scenario 2 assumes that some people opt to use self-sampling instead of routine screening but there is no increase in the overall uptake. Scenario 4 assumes that there is some conversion to self-sampling in current attenders and additional uptake of self-sampling in non-attenders (non-attenders refers to people who at present, given only the option to have routine screening but do not screen) [17].

In Scenario 3, self-sampling is offered only to non-attenders with a range of uptake informed by published studies. Scenarios 3d and 3h take into account the ‘nudge effect’, in which non-attenders being invited to self-sample choose to attend for routine screening [18]. While very high conversion rates to self-sampling are considered unlikely for vaginal self-sampling [18], recent studies indicate that preference for urine self-sampling could result in higher conversion rates and uptake for urine sampling, [19–25] and as such are considered in Scenario 2c and 4c.

**Supplementary Table 13. Scenarios used to calculate overall screening costs**

| Scenario | | Description | Clinician-collected (%) | Self-sample (%) | Clinician-collected (n) | Self-sample (n) | Overall uptake (%) | Reference |
|---|---|---|---|---|---|---|---|---|
| Replace one form of testing with another - no change in overall uptake | | | | | | | | |
| SoC | - | Only routine screening offered | 72.0% | 0.0% | 2,908,787 | 0 | 72.0% | Supplementary Table 7 |
| 1a | FV urine | Only self-sampling offered | 0.0% | 72.0% | 0 | 2,908,787 | 72.0% | Assumption |
| 1b | Vaginal swab | Only self-sampling offered | 0.0% | 72.0% | 0 | 2,908,787 | 72.0% | Assumption |
| Self-sampling is offered as an option to all - no change in uptake in non-responders | | | | | | | | |
| 2a | FV urine | Low conversion (25%) to self-sampling | 54.0% | 18.0% | 2,181,590 | 727,197 | 72.0% | Assumption |
| 2b | FV urine | Medium conversion (50%) to self-sampling | 36.0% | 36.0% | 1,454,394 | 1,454,394 | 72.0% | [26] |
| 2c | FV urine | High conversion (75%) to self-sampling | 18.0% | 54.0% | 727,197 | 2,181,590 | 72.0% | Assumption |
| 2d | Vaginal swab | Low conversion (25%) to self-sampling | 54.0% | 18.0% | 2,181,590 | 727,197 | 72.0% | Assumption |
| 2e | Vaginal swab | Medium conversion (50%) to self-sampling | 36.0% | 36.0% | 1,454,394 | 1,454,394 | 72.0% | [26] |
| Self-sampling only offered to those who do not currently screen (non-responders) | | | | | | | | |
| 3a | FV urine | Low uptake of self-sampling (8%) | 72.0% | 2.2% | 2,908,787 | 90,496 | 74.2% | [27] |
| 3b | FV urine | Medium uptake of self-sampling (15%) | 72.0% | 4.2% | 2,908,787 | 169,679 | 76.2% | [17] |
| 3c | FV urine | High uptake of self-sampling (20%) | 72.0% | 5.6% | 2,908,787 | 226,239 | 77.6% | [18] |
| 3d | FV urine | High uptake of self-sampling (20%) plus 10% additional uptake of smear test | 74.8% | 5.6% | 3,021,907 | 226,239 | 80.4% | [18] |

| | | | | | | | | |
|---|---|---|---|---|---|---|---|---|
| 3e | Vaginal swab | Low uptake of self-sampling (8%) | 72.0% | 2.2% | 2,908,787 | 90,496 | 74.2% | [27] |
| 3f | Vaginal swab | Medium uptake of self-sampling (15%) | 72.0% | 4.2% | 2,908,787 | 169,679 | 76.2% | [17] |
| 3g | Vaginal swab | High uptake of self-sampling (20%) | 72.0% | 5.6% | 2,908,787 | 226,239 | 77.6% | [18] |
| 3h | Vaginal swab | High uptake of self-sampling (20%) plus 10% additional uptake of routine screening | 74.8% | 5.6% | 3,021,907 | 226,239 | 80.4% | [18] |
| Self-sampling offered as an option to all – resulting in 15% (medium) uptake in self-sampling in non-responders | | | | | | | | |
| 4a | FV urine | Low conversion (25%) to self-sampling in those who currently screen | 54% | 22% | 2,181,590 | 896,876 | 76.2% | Assumption and [17,26] |
| 4b | FV urine | Medium conversion (50%) to self-sampling in those who currently screen | 36% | 40% | 1,454,394 | 1,624,073 | 76.2% | |
| 4c | FV urine | High conversion (75%) to self-sampling in those who currently screen | 18% | 58% | 727,197 | 2,351,270 | 76.2% | |
| 4d | Vaginal swab | Low conversion (25%) to self-sampling in those who currently screen | 54% | 22% | 2,181,590 | 896,876 | 76.2% | |
| 4e | Vaginal swab | Medium conversion (50%) to self-sampling in those who currently screen | 36% | 40% | 1,454,394 | 1,624,073 | 76.2% | |

FV, first void; SoC, standard of care.

**Supplementary Table 14. Break-down of calculations in scenarios.**

| Scenario | | Description | Total cost | | Complete screens (n) | | Colposcopies (n) | |
|---|---|---|---|---|---|---|---|---|
| | | | Routine screening | Self-sample | Routine screening | Self-sample | Routine screening | Self-sample |
| Replace one form of testing with another - no change in overall uptake | | | | | | | | |
| SoC | - | Only routine screening offered | £155,658,155 | £0 | 2,739,987 | - | 96,804 | - |
| 1a | FV urine | Only self-sampling offered | £0 | £104,990,567 | - | 2,722,134 | - | 86,080 |
| 1b | Vaginal swab | Only self-sampling offered | £0 | £109,890,829 | - | 2,722,134 | - | 86,080 |
| Self-sampling is offered as an option to all - no change in uptake in non-responders | | | | | | | | |
| 2a | FV urine | Low conversion (25%) to self-sampling | £116,743,616 | £26,247,642 | 2,054,990 | 680,533 | 72,603 | 21,520 |
| 2b | FV urine | Medium conversion (50%) to self-sampling | £77,829,078 | £52,495,284 | 1,369,994 | 1,361,067 | 48,402 | 43,040 |
| 2c | FV urine | High conversion (75%) to self-sampling | £38,914,539 | £78,742,926 | 684,997 | 2,041,600 | 24,201 | 64,560 |
| 2d | Vaginal swab | Low conversion (25%) to self-sampling | £116,743,616 | £27,472,707 | 2,054,990 | 680,533 | 72,603 | 21,520 |
| 2e | Vaginal swab | Medium conversion (50%) to self-sampling | £77,829,078 | £54,945,414 | 1,369,994 | 1,361,067 | 48,402 | 43,040 |
| Self-sampling only offered to those who do not currently screen (non-responders) | | | | | | | | |
| 3a | FV urine | Low uptake of self-sampling (8%) | £155,658,155 | £3,266,373 | 2,739,987 | 84,689 | 96,804 | 2,678 |
| 3b | FV urine | Medium uptake of self-sampling (15%) | £155,658,155 | £6,124,450 | 2,739,987 | 158,791 | 96,804 | 5,021 |
| 3c | FV urine | High uptake of self-sampling (20%) | £155,658,155 | £8,165,933 | 2,739,987 | 211,722 | 96,804 | 6,695 |
| 3d | FV urine | High uptake of self-sampling (20%) plus 10% additional uptake of routine screening | £161,711,528 | £8,165,933 | 2,846,542 | 211,722 | 100,568 | 6,695 |

| | | | | | | | | |
|---|---|---|---|---|---|---|---|---|
| 3e | Vaginal swab | Low uptake of self-sampling (8%) | £155,658,155 | £3,418,826 | 2,739,987 | 84,689 | 96,804 | 2,678 |
| 3f | Vaginal swab | Medium uptake of self-sampling (15%) | £155,658,155 | £6,410,298 | 2,739,987 | 158,791 | 96,804 | 5,021 |
| 3g | Vaginal swab | High uptake of self-sampling (20%) | £155,658,155 | £8,547,064 | 2,739,987 | 211,722 | 96,804 | 6,695 |
| 3h | Vaginal swab | High uptake of self-sampling (20%) plus 10% additional uptake of routine screening | £161,711,528 | £8,547,064 | 2,846,542 | 211,722 | 100,568 | 6,695 |
| Self-sampling offered as an option to all – resulting in 15% (medium) uptake in self-sampling in non-responders | | | | | | | | |
| 4a | FV urine | Low conversion (25%) to self-sampling in those who currently screen | £116,743,616 | £32,372,092 | 2,054,990 | 839,325 | 72,603 | 26,541 |
| 4b | FV urine | Medium conversion (50%) to self-sampling in those who currently screen | £77,829,078 | £58,619,733 | 1,369,994 | 1,519,858 | 48,402 | 48,062 |
| 4c | FV urine | High conversion (75%) to self-sampling in those who currently screen | £38,914,539 | £84,867,375 | 684,997 | 2,200,392 | 24,201 | 69,582 |
| 4d | Vaginal swab | Low conversion (25%) to self-sampling in those who currently screen | £116,743,616 | £33,883,006 | 2,054,990 | 839,325 | 72,603 | 26,541 |
| 4e | Vaginal swab | Medium conversion (50%) to self-sampling in those who currently screen | £77,829,078 | £61,355,713 | 1,369,994 | 1,519,858 | 48,402 | 48,062 |

FV, first void; SoC, standard of care.

**Section 4: Calculating cost per complete screen given low and high values for HR HPV positivity**

Data from an observational study in England on the impact of the bivalent HR-HPV vaccine on cervical screening outcomes was used to inform assumptions on relative HR-HPV positivity reduction in a future vaccinated cohort [28] (Supplementary Table 15). The main model was then rerun with the low HR-HPV positivity values at baseline, year 2 and year 3 to calculate the cost per complete screen (Supplementary Table 16).

**Supplementary Table 15. Variables and calculations used to inform the low HR-HPV infection values in scenario analysis**

| Screening outcome | | Percentage | Reference/Comment |
|---|---|---|---|
| Assumed relative HR HPV positivity in a partially vaccinated compared to a non-vaccinated cohort | | | |
| Percentage of women aged 24-25 years positive for HR HPV when 0% of the cohort are vaccinated | a | 33.7% | English study, Table 1, 2013 [28] |
| Percentage of women aged 24-25 years positive for HR HPV when 55% of the cohort are vaccinated | b | 25.9% | English study, Table 1, 2018 [28] |
| Relative HR HPV positivity | c | 76.9% | Calculated variable (b/a). |
| Base case HR HPV positivity | | | |
| Percentage HR-HPV positive at baseline | d | 11.5% | English pilot study [2] Calculated in Supplementary Table 9. |
| Percentage HR-HPV positive at year 2 (12-month recall) | e | 56.5% | English pilot study [2] Calculated in Supplementary Table 10. |
| Percentage HR-HPV positive at year 3 (24-month recall) | f | 65.8% | English pilot study [2] Calculated in Supplementary Table 10. |
| Assumed lower (reduced) HR HPV positivity in a partially vaccinated cohort | | | |
| Low value percentage HR-HPV positive at baseline | | 8.9% | Calculated variable (d*c). |
| Low value percentage HR-HPV positive at year 2 (12-month recall) | | 43.4% | Calculated variable (e*c). |
| Low value percentage HR-HPV positive at year 3 (24-month recall) | | 50.6% | Calculated variable (f*c). |

**Supplementary Table 16. Cost per complete screen - results from the scenario analysis using low values for HR HPV infection**

| Sampling strategy | Total screening cost | Number of complete screens | Cost per complete screen[1] |
|---|---|---|---|
| Clinician-collected cervical sample | £363,607 | 6,897 | £52.72 |
| FV urine self-sample | £236,312 | 6,863 | £34.43 |
| Vaginal self-sample | £248,257 | 6,863 | £36.17 |

[1] Calculated as the total cost of screening divided by the number of complete screens.

2 Rebolj M, Mathews CS, Pesola F, *et al.* Age-specific outcomes from the first round of HPV screening in unvaccinated women: Observational study from the English cervical screening pilot. *BJOG Int J Obstet Gynaecol*;**n/a**. doi:10.1111/1471-0528.17058

3 NHS England. Cervical Screening Programme England - 2020-21 - Data Tables. NHS Digit. https://digital.nhs.uk/data-and-information/publications/statistical/cervical-screening-annual/england---2020-21 (accessed 19 Jan 2022).

4 NHS England. 2021/22 Annex A: The national tariff workbook. 2021.https://www.england.nhs.uk/publication/national-tariff-payment-system-documents-annexes-and-supporting-documents/ (accessed 20 Mar 2022).

5 Moss S, Gray A, Marteau T, *et al.* Evaluation of HPV/LBC Cervical Screening Pilot Studies Report to the Department of Health. 2004. http://www.cancerscreening.nhs.uk/cervical/evaluation-hpv-2006feb.pdf

6 PSSRU. Unit Costs of Health and Social Care 2021 | PSSRU. 2021. https://www.pssru.ac.uk/project-pages/unit-costs/unit-costs-of-health-and-social-care-2021/ (accessed 20 Mar 2022).

7 Royal Mail. Royal Mail Business Price Guide January 2022. 2022.https://www.royalmail.com/sites/royalmail.com/files/2021-12/royal-mail-business-price-guide-january-2022-v1.pdf (accessed 23 Feb 2022).

8 Royal Mail. Royal Mail Response Services Rate Card January 2022. 2022.https://www.royalmail.com/sites/royalmail.com/files/2022-01/response-services-rate-card-january-2022.pdf (accessed 23 Feb 2022).

9 Kitchener HC, Gittins M, Rivero-Arias O, *et al.* A cluster randomised trial of strategies to increase cervical screening uptake at first invitation (STRATEGIC). *Health Technol Assess Winch Engl* 2016;**20**:1–138. doi:10.3310/hta20680

10 Flocked swabs. VWR. https://uk.vwr.com/store/product/12272349/flocked-swabs (accessed 28 Mar 2022).

11 Copan Diagnostics Nylon-Flocked Dry Swabs in Tubes - Cell Culture Utensils, Applicators and Swabs. https://www.fishersci.com/shop/products/copan-diagnostics-nylon-flocked-dry-swabs-tubes-7/p-3688222 (accessed 28 Mar 2022).

12 Copan Diagnostics 552C - Floqswab Swab Regular 80mm 1000/Ca - CIA Medical. https://www.ciamedical.com/copan-diagnostics-552c-case-floqswab-swab-regular-80mm-1000-ca (accessed 28 Mar 2022).

13 Sterilin Flocked Swab Regular in Tube 552C | at Zoro. Zoro Tools MRO Supplier. https://www.zoro.co.uk/shop/laboratory-equipment/test-tubes/flocked-swab-regular-in-tube/p/ZT1251115S (accessed 28 Mar 2022).

14 Legood R, Sadique Z, Patnick J, *et al.* Cost and logistics of alternative roll-out options for implementing human papillomavirus testing as a triage in cervical screening: results of the sentinel sites study. *Br J Cancer* 2012;**107**:1574–9. doi:10.1038/bjc.2012.435

15 PreservCyt Solution, 20ml, pack/250. https://supply.unicef.org/s0001682.html (accessed 28 Mar 2022).

16 Cervical Screening (Annual). NHS Digit. https://digital.nhs.uk/data-and-information/publications/statistical/cervical-screening-annual (accessed 30 Mar 2022).

17 Lefeuvre C, Pivert A, Guillou-Guillemette HL, *et al.* Urinary HPV DNA testing as a tool for cervical cancer screening in women who are reluctant to have a Pap smear in France. *J Infect* 2020;**81**:248–54. doi:10.1016/j.jinf.2020.05.006

18 Lam JUH, Rebolj M, Møller Ejegod D, *et al.* Human papillomavirus self-sampling for screening nonattenders: Opt-in pilot implementation with electronic communication platforms. *Int J Cancer* 2017;**140**:2212–9. doi:10.1002/ijc.30647

19 Williams D, Davies M, Fiander A, *et al.* Women's perspectives on human papillomavirus self-sampling in the context of the UK cervical screening programme. *Health Expect Int J Public Particip Health Care Health Policy* 2017;**20**:1031–40. doi:10.1111/hex.12544

20 Sargent A., Fletcher S., Bray K., *et al.* Cross-sectional study of HPV testing in self-sampled urine and comparison with matched vaginal and cervical samples in women attending colposcopy for

the management of abnormal cervical screening. *BMJ Open* 2019;**9**:e025388. doi:10.1136/bmjopen-2018-025388

21 Lozar T, Nagvekar R, Rohrer C, *et al.* Cervical Cancer Screening Postpandemic: Self-Sampling Opportunities to Accelerate the Elimination of Cervical Cancer. *Int J Womens Health* 2021;**13**:841–59. doi:10.2147/IJWH.S288376

22 Tranberg M, Jensen JS, Bech BH, *et al.* Urine collection in cervical cancer screening - analytical comparison of two HPV DNA assays. *BMC Infect Dis* 2020;**20**:926. doi:10.1186/s12879-020-05663-7

23 De Pauw H, Donders G, Weyers S, *et al.* Cervical cancer screening using HPV tests on self-samples: attitudes and preferences of women participating in the VALHUDES study. *Arch Public Health* 2021;**79**:155. doi:10.1186/s13690-021-00667-4

24 Schaafsma M, van den Helder R, Bleeker MCG, *et al.* Experiences and preferences towards collecting a urine and cervicovaginal self-sample among women attending a colposcopy clinic. *Prev Med Rep* 2022;**26**:101749. doi:10.1016/j.pmedr.2022.101749

25 Ørnskov D, Jochumsen K, Steiner PH, *et al.* Clinical performance and acceptability of self-collected vaginal and urine samples compared with clinician-taken cervical samples for HPV testing among women referred  for colposcopy. A cross-sectional study. *BMJ Open* 2021;**11**:e041512. doi:10.1136/bmjopen-2020-041512

26 Drysdale H, Marlow LA, Lim A, *et al*. Self-sampling for cervical screening offered at the point of invitation: A cross-sectional study of preferences in England. J Med Screening 2022 9691413221092246 Online ahead of print. doi: 10.1177/09691413221092246

27 Arbyn M, Smith SB, Temin S, *et al.* Detecting cervical precancer and reaching underscreened women by using HPV testing on self samples: updated meta-analyses. *BMJ* 2018;**363**:k4823. doi:10.1136/bmj.k4823

28 Rebolj M, Pesola F, Mathews C, *et al.* The impact of catch-up bivalent human papillomavirus vaccination on cervical screening outcomes: an observational study from the English HPV primary screening pilot. *Br J Cancer* Published Online First: 26 March 2022. doi:10.1038/s41416-022-01791-w
